# REduction of the lifting Load Among logistics workers through a passive back eXoskeleton. Protocol of the RELAX project, an 18-month in-field controlled intervention study

**DOI:** 10.64898/2026.05.21.26353770

**Authors:** Lasse Schrøder Jakobsen, Sebastian Skals, David Høyrup Christiansen, Jan Sørensen, Charles Pontonnier, Pascal Madeleine

## Abstract

**Background:** Occupational exoskeletons are used to reduce physical workload and prevent work-related musculoskeletal disorders in physically demanding jobs. Although laboratory studies demonstrate reduced muscle load during simulated manual work tasks, evidence from long-term, real-world implementations remains very limited. The RELAX project aims to investigate the long-term effects of a passive back-support exoskeleton (BSE) during manual order-picking work in a Danish warehouse, focusing on health and socio-economic outcomes.

**Methods:** This 18-month controlled in-field intervention study compares outcomes at two warehouse departments: one where workers use a passive BSE and a control group where workers perform work tasks as usual. Approximately 90 full-time workers will be followed during the intervention period with questionnaires, interviews and company-registered performance indicators. Primary outcomes include perceived work intensity and musculoskeletal discomfort, while secondary outcomes include sickness absence, employee turnover, productivity and cost effectiveness.Furthermore, a process evaluation will be conducted based on questionnaires, focus-group interviews, and reported exoskeleton use. Quantitative effects will be analysed using difference-indifference analysis with generalized linear mixed models to account for repeated measures over time. Employee turnover will be analysed using time-to-event analysis, and qualitative focus-group interviews will be analysed using reflexive thematic analysis to explore implementation processes and contextual factors. Cost-effectiveness and return on investment will be assessed by comparing the investment with potential savings in costs and resource use.

**Discussion:** By combining longitudinal quantitative outcomes with qualitative process evaluation, the study seeks to provide ecologically valid evidence on the effectiveness, feasibility and sustainability of occupational exoskeleton implementation. This approach will help clarify whether long-term exoskeleton use improves worker health without compromising productivity and may inform future workplace guidelines and large-scale adoption strategies.

**Trial Registration:** ClinicalTrials.gov identifier: NCT07561112. Registered May 7, 2026.

## Background

Occupational exoskeletons are wearable assistive devices designed to support and relieve the musculoskeletal system during physical labour (1). Over the past decades, research has demonstrated the potential of this technology to reduce physical load across a wide range of manual work tasks (1-3). Consequently, these devices are increasingly considered an attractive solution for preventing worker attrition and work-related musculoskeletal disorders (WMSDs) (4). This is particularly relevant in physically demanding sectors such as logistics, which involve heavy lifting, awkward postures, and repetitive movements (5-7).

Several studies assessing heart rate, muscle activity, and kinematics have documented benefits of back-supporting exoskeletons (BSE) during both static and dynamic work tasks, primarily through reductions in the level of muscle activity (8-10). However, existing literature highlights a significant lack of longitudinal field studies on occupational exoskeletons across different work environments (1,3,11–13). A few studies have recently investigated the long-term effects of industrial use of shoulder- and back-supporting exoskeletons on work intensity and musculoskeletal discomfort (14,15). Nevertheless, there remains a need for comprehensive analyses involving large-scale workplace implementation of exoskeletons.

A recent systematic review listed several documented side effects associated with occupational use of exoskeletons (13). While mild discomfort and restricted mobility are among the most frequently reported issues, the review emphasizes that the effects of long-term use (> 3-months) should be examined to achieve a thorough understanding of how these side effects influence occupational exoskeleton use. Additionally, previous research suggests multiple benefits of training and familiarization, highlighting the importance of investigating not only the short-term effects of exoskeleton use but also the effects of prolonged usage periods (10,16-18).

We have recently conducted a randomized controlled trial to investigate the use of BSE among warehouse workers performing daily order-picking tasks over a 24-week period. We reported positive longitudinal effects, including consistently reduced level of muscle activity and perceived effort during simulated work tasks at pre and post-tests, as well as progressive reductions in perceived work intensity during the intervention. However, no significant reductions in musculoskeletal discomfort were observed (19). As highlighted in several systematic reviews (1-3,20,11-13), large-scale interventions involving a greater number of workers and longer study durations are needed to clarify the long-term effects of exoskeleton use on socioeconomic parameters, such as sick leave, productivity, employee turnover, and musculoskeletal discomfort.

To accommodate the recommendation highlighted in the previous conducted reviews, the present study aims to investigate the effects of 18 months of occupational use of a BSE during manual order-picking tasks in a Danish warehouse. We hypothesize that long-term use of the exoskeleton will reduce perceived work intensity, musculoskeletal discomfort, sick leave, and employee turnover, without negatively affecting productivity. Further, we hypothesize that investment in BSE is cost-effective and offer good return on investment.

## Methods

### Study design

The study is designed as a controlled intervention study. Participants will be enrolled in two experimental groups from two different warehouse departments of similarly sized workforces, respectively, performing daily manual order picking tasks: 1) *the intervention group* performing their daily work tasks using a BSE, and 2) *the control group* performing their daily work tasks as usual with no added assistance. The study is conducted at Dagrofa Logistik A/S in Vejle, Denmark and was designed in collaboration with Aalborg University, RCSI University of Medicine and Health Sciences, Ecole Normale Supérieure de Rennes, The National Research Centre for the Working Environment, and Aarhus University Hospital.

Future reporting of the trial will follow Consolidated Standards of Reporting Trials (CONSORT) guidelines (21) and the template for Intervention Description and Replication (TIDieR) (22). The trial is being conducted according to the Declaration of Helsinki III. The protocol, template informed consent forms, and participant information have been approved by the Ethics Committee of North Denmark Region (LBK nr. 1268). All participants will provide informed consent before enrolment. Before the inclusion of the first participant, the trial will be registered on clinicaltrials.gov (NCT07561112).

### Recruitment

Participants for the two experimental groups will be recruited among workers from two warehouse departments at Dagrofa Logistik’s terminal in Vejle, Denmark. The intervention group (INT) will be recruited from the department handling fruits and vegetables, while the control group (CON) was recruited from the department handling cooled goods. Although the two departments differ in the type of goods handled, their working environment and processes are identical in terms of temperature (5° Celsius) and applied trucks. Potential differences in volume handled (weight and frequency) in the two departments will be reported during the intervention period. Eligible workers will be identified and will receive information about the trial before giving their informed consent to participate.

### Inclusion criteria

Workers aged 18–65 years with full-time employment at Dagrofa Logistik will be eligible for inclusion. Inclusion criteria are: 1) age 18–65 years at enrolment, 2) full-time employment in the current department, and 3) no pain or injuries affecting daily work tasks. Exclusion criteria were: 1) anthropometric characteristics preventing adequate fit of the exoskeleton, 2) part-time employment, and 3) pregnancy or anticipated prolonged absence from work during the study period (standard vacation not included).

If a participating worker from either the INT or CON group transfers to another department within the company during the intervention period for reasons unrelated to the intervention, the participant will be excluded from the study. Such cases will be classified as dropouts and the corresponding data will be reported as lost at follow-up.

### Interventions

The intervention consists of a workplace implementation of passive BSE for manual order-picking tasks in a real-world logistics setting. The intervention is conducted at Dagrofa Logistik and targets the fruit and vegetable department, which is characterized by high physical workload, frequent manual lifting, and low degree of automation. The principal investigator will be present at the site of the intervention 2-3 times a week to manage and facilitate the intervention.

#### The IX Back Air

The IX Back Air (SUITX by Ottobock) is a passive BSE designed to reduce mechanical loading of the lumbar spine during manual material handling tasks involving forward bending and lifting. The device operates without motors, electronics, or external power sources and provides assistance through a mechanical structure that transfers part of the upper-body load to the pelvis and lower extremities. The BSE weighs approximately 3 kg and is worn externally over work clothing. It features adjustable components that allow individual fitting across a range of body sizes and anthropometries (small, medium, large, x-large) making one size fit the most. Assistance is provided primarily during trunk flexion and lifting movements, while the design permits unrestricted walking and upright postures, enabling use in dynamic work environments such as order picking and warehouse logistics^1^.

The BSE provides a maximal support moments of 24.8 Nm during flexion and 19.2 Nm during extension of the trunk (23). The support is automatically engaged during forward bending and load handling and disengaged during upright activities, minimizing interference with normal movement patterns. The IX Back Air is intended for prolonged occupational use and can be donned and doffed quickly, facilitating its integration into regular work routines. Within the present study, the IX Back Air is used as an ergonomic intervention to reduce cumulative biomechanical loading of the lower back during physically demanding tasks.

#### Implementation procedure

To ensure high adoption, usability, and intervention fidelity, the implementation will follow a stepwise, rollout protocol based on prior field experience (18). The implementation will begin with an introductory phase, including on-site kick-off sessions where employees receive information about the purpose of the study, the functionality of the exoskeleton, expected benefits, and potential limitations. These sessions will emphasize voluntary use, user safety, and correct donning and doffing procedures.

During the rollout phase, 5-10 workers will be introduced to the exoskeleton device per week, allowing gradual familiarization and continuous technical and ergonomic support. On-site assistance from trained manufacturer representatives and the principal investigator will be provided throughout the implementation phase to ensure appropriate individual fitting, adjustment to anthropometric characteristics, and task-specific optimization. Subsequently, a five-step familiarization phase will be initiated, consisting of progressive exoskeleton usage from approx. 7h30min of use per workweek during the first step, to full-time usage during the fifth step (18). Full time usage will vary in hours from worker to worker as the BSE is only worn during order picking tasks, while some workers perform other tasks as well during the workday. A full-time workweek is equivalent to 36 hours. The duration of the familiarization period will depend on user feedback and process evaluations. During the intervention period, employees will be encouraged to use the exoskeleton during physically demanding order-picking tasks but are free to disengage the device when not required (e.g., during walking), reflecting realistic work practices (Fig. 1).

**Figure 1:**
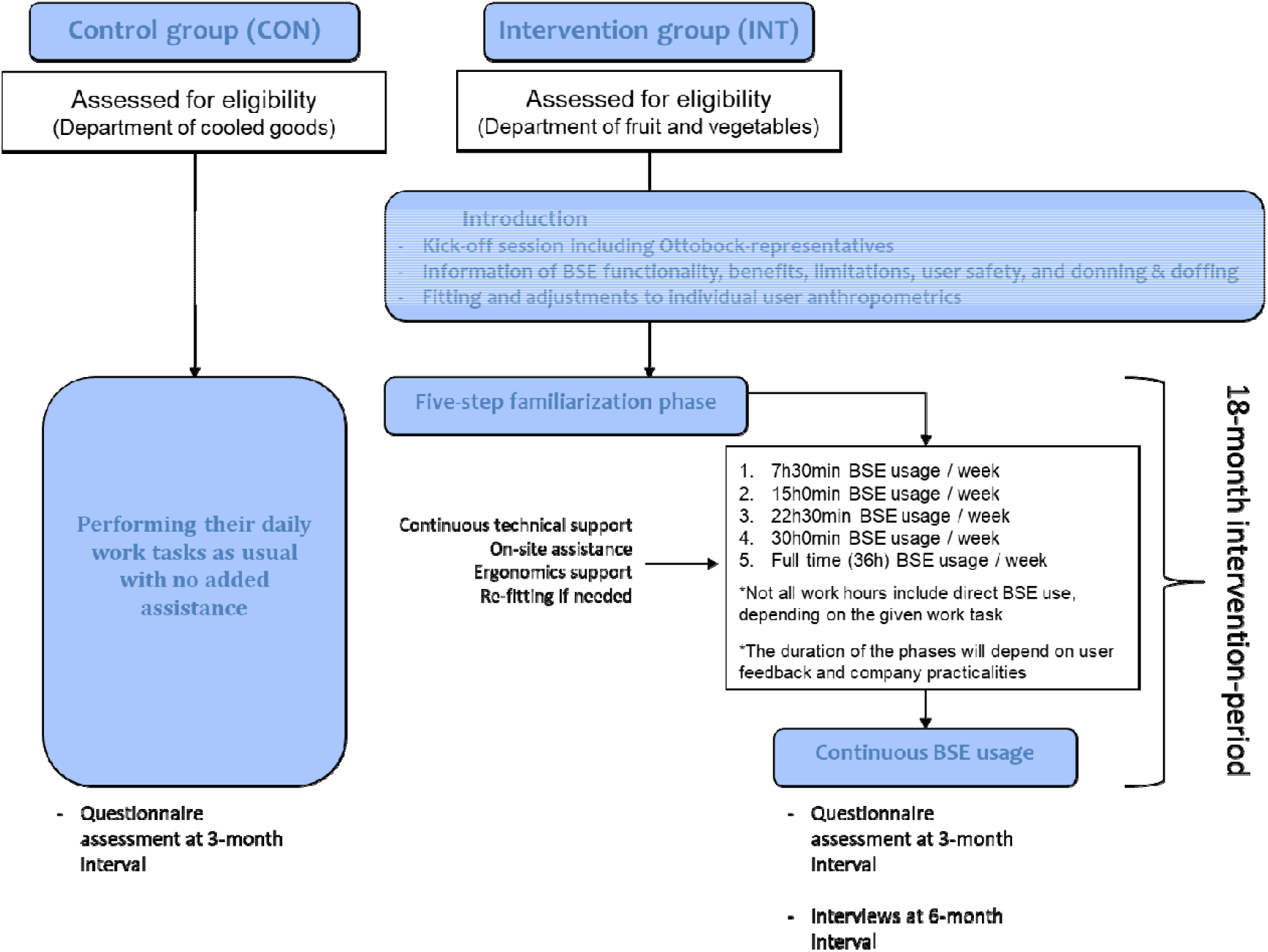
Flowchart of intervention for the control (CON) and intervention group (INT), respectively. This includes the overall introduction of the BSE, the stepwise familiarization phases, and continuous use of the BSE.

### Randomisation and blinding

Randomisation will not be performed due to the study design, which involved recruitment from two separate departments. This approach was chosen as the most feasible solution, given the company’s practical interest in implementing exoskeletons in the fruit and vegetable department. To account for the lack of randomisation, a difference-in-differences (DiD) analysis will be used to compare the causal outcomes between the two groups (24).

No sham or placebo intervention will be used to blind participating workers. However, a blinded interpretation of the statistical analyses will be conducted to minimise interpretative bias (25).

### Outcome variables

The outcome variables are summarised in Table 1.

**Table 1:** Assessments and instruments.

| Outcome | Instrument | Timepoint |
| --- | --- | --- |
| **Primary outcomes** |  |  |
| Perceived work intensity | Two 10-point Likert-scale questions (14). | T0, T3, T6, T9, T12, T15, T18 |
| Musculoskeletal discomfort | Cornell Musculoskeletal Discomfort Questionnaire. | T0, T3, T6, T9, T12, T15, T18 |
| **Secondary outcomes** |  |  |
| Sickness absence | Key performance indicator from the company – reports of sickness absence (number of days) from each individual worker.<br>Questionnaire – *if any, have your recent sickness absence been related to musculoskeletal discomfort?*<br>*Sick days related to musculoskeletal discomfort:*<br>*Sick days related to other factors:* | Weekly<br>T0, T3, T6, T9, T12, T15, T18 |
| Employee turnover | Reports of individual resignations. | Time-to-event |
| Productivity | Key performance indicator from the company – reports of packages handled per hour reported in weekly average. | Weekly |
| Cost-effectiveness and return on investment | Initial BSE and training costs will be quantified.<br>Intervention effects will be monetised from company perspectives. Cost-effectiveness will be expressed relative to avoided sickness absence, | T0, T18 |
|  | reduced turnover, and productivity gains, summarised as return on investment. |  |
| **Other outcomes** |  |  |
| Exoskeleton evaluation / User acceptance | Six 10-point Likert-scale questions and three open-ended questions (14). Including focus-group interview evaluations for elaboration. | T0, T3, T6, T9, T12, T15, T18 |
| Process evaluation | Focus-group interviews with workers from the intervention group and stakeholders, respectively. | T0, T6, T12, T18 |
| Exoskeleton use | Monitored daily by the company using operational logs and reported in weekly average. | Weekly |
| *T0; baseline, T3; baseline +3 month, T6; baseline +6 months, T9; baseline+9 months, T12; baseline +12months, T15; baseline+15 months, T18; baseline +18 months* |  |  |

#### Descriptive

At baseline, the following worker characteristics will be collected: age, sex, work experience (length of employment), body mass, height, and body mass index (BMI) will be calculated. In addition, participants will complete the Cornell Musculoskeletal Discomfort Questionnaire (CMDQ) to report musculoskeletal symptoms (discomfort and pain) experienced during the past workweek (26).

#### Primary outcomes

The primary outcomes cover the effects of exoskeleton use on health-related measures and include:

1. **Perceived work intensity** assessed using two 10-point Likert-scale questions on 1) how much they exert themselves during a shift and 2) how exhausted they feel at the end of a shift (see appendix 1).
2. **Musculoskeletal discomfort** assessed using the CMDQ, completed by workers as part of the questionnaire battery (26). Analyses will primarily focus on reports of back discomfort (see appendix 1).

#### Secondary outcomes

The secondary outcomes cover the effect of exoskeleton use on organizational and company related measures and include:

1. Sickness absence Sickness absence is measured as the number of days of sick leave per month as reported by the company. To assess whether sick leave was related to musculoskeletal issues, the questionnaire will include the question: *“Have you had any sick leave related to musculoskeletal discomfort within the last month?* *Sick days related to musculoskeletal discomfort:______* *Sick days related to other factors: ______”* (see appendix 1).
2. Employee turnover Employee turnover is measured as the number of monthly resignations in the respective department as reported by the company.
3. Productivity Productivity is a key performance indicator monitored by the company reported in packages per hour for each individual worker. In this study productivity will be reported as a weekly average.
4. Cost-effectiveness and return on investment The initial investment in BSE and implementation will be quantified to assess the cost and resource of the intervention. The consequences of the intervention in terms of reduced sickness absence and increase productivity will be quantified and monetarised from a company view. Cost-effectiveness will be evaluated by comparing intervention costs with the combined economic benefits, including avoided costs from reduced sickness absence and worker turnover, as well as increased earnings due to productivity gains. These measures will be accumulated and summarised to a single measure of return on investment.

#### Other outcomes

The remaining outcomes cover exoskeleton and process evaluations and include:

1. Exoskeleton evaluation / User acceptance Evaluations of the exoskeleton will be conducted using six 10-point Liker-Scale questions for assessment of comfort, thermal comfort, balance, range of motion, safety, and performance. Additionally, the questionnaire will include three open-ended questions on what they like most and least about the BSE, and what they would like to change about the BSE if possible. The questions are adapted from Kim et al., 2021 (14), and will additionally form the basis of the focus-group interviews for further elaboration (see appendix 1).
2. Process evaluation A process evaluation will be conducted to support interpretation of the intervention outcomes and to document implementation fidelity, user acceptance, and contextual factors influencing exoskeleton use. Process evaluation data will be collected every 6^th^ month throughout the 18-month intervention period through repeated focus-group interviews with workers from the intervention group and relevant stakeholders. The interviews will explore experiences with BSE use, integration into daily work routines, perceived benefits and drawbacks, and organisational factors affecting implementation.
3. Exoskeleton use During the intervention period, workers will report daily exoskeleton use using operational logs. Specifically, usage will be recorded through a stamp system each time the exoskeleton is used. In this study exoskeleton use will be reported as a weekly average.

### Sample size

The sample size is limited by the total pool of workers employed at the two departments. Both departments have a workforce of 50-55 full-time employees at any given time. All workers will be assessed for eligibility, giving an expected total sample of approximately 90 employees, corresponding to 81 participants after an anticipated 10% dropout at follow-up. Power analyses were conducted using an F-test for repeated measures within–between interaction, corresponding to the primary *group x time* effect evaluated in the study.

For perceived work intensity, an a priori power analysis was performed using effect size estimates from the previously conducted 24-week intervention study, which reported significant *time x group* interactions for perceived work intensity (19). To avoid overestimating the effect based on the small sample of the previous study, a conservative effect size of f = 0.30 was applied. Assuming α = 0.05, power = 0.80, two groups, seven repeated measurements, a correlation among repeated measures of 0.5, and a non-sphericity correction of ε = 0.75, the required total sample size was 16 participants. This indicates that the expected study population is sufficient to detect an intervention effect of this magnitude.

For musculoskeletal discomfort, the previous study did not report a significant *time x group* interaction, and therefore no reliable effect size could be derived from those results (19). Consequently, a sensitivity analysis was conducted using the expected final sample size of 81 participants (assuming 10% drop-out). Under the same statistical assumptions, the study would be able to detect a minimum effect size of 0.12, corresponding to a small interaction effect.

Overall, these analyses indicate that the planned sample size should provide adequate statistical power to detect small-to-moderate longitudinal intervention effects in the questionnaire outcomes.

If less than 90 employees (45 in each group) are enrolled at baseline, a rolling inclusion will be applied, enrolling newly employed workers until this is reached.

### Statistical analysis

All statistical analyses will be conducted according to a predefined analysis plan. Analyses will primarily follow an intention-to-treat principle, including all participants enrolled at baseline (and later if needed to reach a sample size of 45 in each group, respectively), regardless of adherence to exoskeleton use. Descriptive statistics will be used to summarise baseline characteristics and outcome distributions for the intervention and control groups.

Because participants are recruited from two separate departments without randomisation, baseline differences between the intervention and control groups may occur. Baseline characteristics including age, anthropometric variables (height, body mass and BMI), work experience, and musculoskeletal discomfort will be summarised descriptively and compared between groups. If relevant imbalances are identified, these variables will be included as covariates in the mixed-effects models to account for potential confounding and to ensure that estimated intervention effects are not driven by pre-existing group differences.

Prior to inferential analyses, assumptions underlying parametric models will be evaluated. Normality of continuous outcomes will be assessed using visual inspection of histograms and Q–Q plots, supplemented by Shapiro–Wilk tests where appropriate. Homogeneity of variance between groups at baseline will be examined using Levene’s test.

Given the non-randomised allocation of participants to groups, intervention effects will be evaluated using a DiD approach to compare the causal changes in outcomes over time between the intervention and control departments (24). This method accounts for baseline differences between groups and for common temporal trends unrelated to the intervention. The primary parameter of interest will be the *group x time* interaction, representing the estimated effect of exoskeleton implementation beyond secular changes.

Intervention effects (Musculoskeletal discomfort, Perceived Work Intensity, Sickness absence, and Productivity) will be evaluated using generalized linear mixed models (GLMMs) to account for the longitudinal and clustered structure of the data. Repeated measurements (questionnaire and KPI data) are within subject factors, while group (intervention or control) are between subject factors. All models will include a random intercept for participant to account for within-subject correlation across time.

Fixed effects will include group (intervention vs control), time (categorical month), and the *group x time* interaction. The *group x time* interaction represents the DiD estimate and is interpreted as the effect of exoskeleton implementation beyond secular time trends and baseline differences between departments.

Continuous outcomes (musculoskeletal discomfort, perceived work intensity and productivity) will be analysed using linear mixed models, while count outcomes (number of sick-leave days) using negative binomial mixed models. An additional analysis on sickness absence will be conducted, only assessing sick-days related to musculoskeletal discomfort based on the binary questionnaire – *‘if any, have your recent sick absence been related to musculoskeletal discomfort?’*.

Model fit will be assessed using residual checks and comparison of alternative correlation structures. Results will be presented as estimated group means over time with 95% confidence intervals.

Employee turnover will be analysed using time to event methods. The outcome will be defined as the time from inclusion until an event, where the event is voluntary resignation from employment. A time variable will represent the duration (in months) from individual baseline (T0) until resignation or last observation. A status variable will be created indicating whether the event occurred (1 = resignation) or the observation was censored (0 = still employed or lost to follow-up for other reasons).

Participants who remain employed at the end of the study period or who leave for reasons unrelated to resignation (e.g., long-term leave or end of study follow-up) will be treated as censored cases at their last known observation time.

Kaplan–Meier curves will be generated to estimate the survival function (probability of remaining employed) over time for intervention and control groups, and differences between groups will be assessed using the log-rank test. To quantify the relative risk of turnover, a Cox proportional hazards regression model will be fitted with group as the main predictor. Hazard ratios (HR) with 95% confidence intervals will be reported. The proportional hazards assumption will be evaluated graphically using log–log survival plots and by testing time-dependent effects.

Qualitative data obtained from the repeated focus-group interviews will be analysed using reflexive thematic analysis to explore workers’ and stakeholders’ experiences with the exoskeleton implementation and to support interpretation of quantitative findings. All interviews will be audio-recorded and transcribed prior to analysis.

The analysis will follow the six phases described by Braun and Clarke (27): (1) familiarization with the data through repeated reading of transcripts, (2) initial inductive coding of meaningful text segments, (3) organisation of codes into candidate themes, (4) review and refinement of themes in relation to the full dataset, (5) definition and naming of themes, and (6) reporting of themes with illustrative quotations. Coding will primarily follow an inductive approach, while also allowing deductive consideration of implementation aspects such as usability, acceptance, barriers, and facilitators.

The qualitative findings will be integrated with quantitative outcomes in a mixed-methods framework to contextualise observed effects and explain mechanisms underlying adoption, adherence, and potential unintended consequences of the intervention.

All quantitative analyses will be conducted using the statistical analysis software SPSS version 26 (IBM crop., Armonk, NY, USA). P-values below 0.05 will be considered as significant.

### Data monitoring and quality assurance

All data will be stored in accordance with the Danish Personal Data Protection Act and other applicable Danish legislation. Questionnaires will be translated to the native tongue of the workers using published guidelines (28) and will be collected in *Microsoft Forms* during outcome assessments. All datasets will be fully anonymised and will not contain any personal identifying information. Personal data linking participants to study identifiers will be stored securely by the principal investigator and will not be shared.

All primary and secondary outcome analyses will be performed by the principal investigator. To minimise the risk of interpretative bias, a blinded interpretation procedure will be conducted by members of the research team, excluding the principal investigator. Prior to dissemination, the principal investigator will code the study groups as *Group A* and *Group B* and provide the statistical results to the blinded authors. The blinded authors will independently formulate two alternative interpretations of the findings: one assuming Group A as the intervention group and Group B as the control group, and one with the allocation reversed. These interpretations will be preregistered in a document entitled *“The RELAX-trial: Blinded data analyses interpretation statement*.*”* After registration, group allocation will be revealed, and the correct interpretation will be selected as the basis for the discussion of the primary publication (25). To maintain non-disclosure between the author groups, this process will include only measures collected for both experimental groups.These measures include perceived work intensity, musculoskeletal discomfort, sickness absence, employee turnover, and productivity.

## Discussion

This protocol describes the RELAX project: a large-scale, long-term controlled intervention study investigating the effects of occupational use of a passive back-support exoskeleton during manual order-picking tasks in a real-world warehouse setting. The study addresses a critical gap in the existing literature by moving beyond short-term laboratory evaluations and examining longitudinal health and socio-economic outcomes associated with exoskeleton implementation (1,3,7,11-13,20).

A major strength of the study is its in-field design, which allows assessment of BSE use under authentic working conditions without altering existing work organisation, productivity targets, or task allocation. This will enhance ecological validity and ensures that observed effects are relevant to real-world implementation. The extended intervention period of 18 months will enable evaluation of both potential benefits and unintended consequences associated with prolonged use, including adaptation, adherence, and user acceptance over time.

The study will incorporate multiple outcomes, including perceived work intensity, musculoskeletal discomfort, sickness absence, employee turnover, productivity, and process-related measures. This comprehensive approach reflects the multifactorial aspects of work-related musculoskeletal health and acknowledges that successful implementation of assistive technologies depends not only on biomechanical effects but also on organisational and socio-economic factors (29). The combination of quantitative outcomes with qualitative process evaluations will also provide additional insight into mechanisms underlying observed effects and potential barriers to large-scale adoption.

Several methodological limitations should be acknowledged. First, the lack of randomisation may introduce residual confounding due to unmeasured differences between departments. However, the causal inference from the application of DiD analyses mitigates this limitation by controlling for baseline differences and common temporal trends (24). Second, blinding of participants is not feasible in an intervention involving wearable assistive devices, although blinded interpretation of the statistical analyses is used to reduce interpretative bias (25). This could also influence the assessment of workers in the control group from their information of the experience of the intervention group workers. Third, outcomes such as musculoskeletal discomfort and perceived work intensity rely on self-reported measures, which may be influenced by expectations or reporting bias (30).

Despite these limitations, the RELAX project is well positioned to generate robust evidence on the long-term effects of BSE in physically demanding logistics work. If positive effects on worker well-being are observed without adverse impacts on productivity, the findings may inform evidence-based decisions regarding occupational exoskeleton implementation and contribute to the development of guidelines for their safe and effective use. Conversely, identification of limited benefits or unintended side effects will be equally valuable for guiding future research, device development, and workplace policy.

In conclusion, this study will contribute with important and robust analyses to enhance the understanding of the real-world effectiveness, feasibility, and sustainability of occupational exoskeletons as an ergonomic intervention in the logistics sector.

## Data Availability

All data produced in the present study are available upon reasonable request to the authors

## Footnotes

1 IX BACK AIR exoskeleton | SUITX

