## Appendix 1 for "REduction of the lifting Load Among logistics workers through a passive back eXoskeleton. Protocol of the RELAX project, an 18-month in-field controlled intervention study"

### Appendix 1. Questionnaire Battery

**If any, have your recent sick absence been related to musculoskeletal discomfort?**

**Sick days related to musculoskeletal discomfort:**

**Sick days related to other factors:**

#### Answer the questions be marking 0-10:

**What is your perception of the overall fit and comfort of the exoskeleton when performing your job?**

**0 = No discomfort 10 = Most discomfort**

| | | | | | | | | | | |
|---|---|---|---|---|---|---|---|---|---|---|
| 0 | 1 | 2 | 3 | 4 | 5 | 6 | 7 | 8 | 9 | 10 |

**What is your perception of the thermal comfort typically (and/or feelings of sweatiness) when using the exoskeleton?**

**0 = No discomfort 10 = Most discomfort**

| | | | | | | | | | | |
|---|---|---|---|---|---|---|---|---|---|---|
| 0 | 1 | 2 | 3 | 4 | 5 | 6 | 7 | 8 | 9 | 10 |

**What is your perception of balance (or any sense of imbalance) while using the exoskeleton?**

**0 = perfectly balanced 10 = Off balanced**

| | | | | | | | | | | |
|---|---|---|---|---|---|---|---|---|---|---|
| 0 | 1 | 2 | 3 | 4 | 5 | 6 | 7 | 8 | 9 | 10 |

**Do you feel that your range of motion was at all limited while using the exoskeleton?**

**0 = No limitation 10 = Extremely limiting**

| | | | | | | | | | | |
|---|---|---|---|---|---|---|---|---|---|---|
| 0 | 1 | 2 | 3 | 4 | 5 | 6 | 7 | 8 | 9 | 10 |

**When using the exoskeleton to perform your job, how do you think this affected your overall safety?**

**0 = Substantially less safe 5 = No difference 10 = Substantially safer**

| | | | | | | | | | | |
|---|---|---|---|---|---|---|---|---|---|---|
| 0 | 1 | 2 | 3 | 4 | 5 | 6 | 7 | 8 | 9 | 10 |

**Overall, does using the exoskeleton positively or negatively affect your performance?**

**0 = Substantially worse 5 = No difference 10 = Substantially better**

| | | | | | | | | | | |
|---|---|---|---|---|---|---|---|---|---|---|
| 0 | 1 | 2 | 3 | 4 | 5 | 6 | 7 | 8 | 9 | 10 |

#### Disagree or agree by marking 0-10:

**When I work, I really exert myself to the fullest.**

**0 = Strongly disagree 10 = Strongly agree**

| 0 | 1 | 2 | 3 | 4 | 5 | 6 | 7 | 8 | 9 | 10 |
|---|---|---|---|---|---|---|---|---|---|---|

**I feel exhausted at the end of a shift.**

**0 = Strongly disagree 10 =Strongly agree**

| 0 | 1 | 2 | 3 | 4 | 5 | 6 | 7 | 8 | 9 | 10 |
|---|---|---|---|---|---|---|---|---|---|---|

#### Answer the questions:

**What do you most like about the exoskeleton?**

**Answer:**

**What do you least like about the exoskeleton?**

**Answer:**

**If you could change anything about the exoskeleton, what would you change?**

**Answer:**

The diagram below shows the approximate position of the body parts referred to in the questionnaire. Please answer by marking the appropriate box.

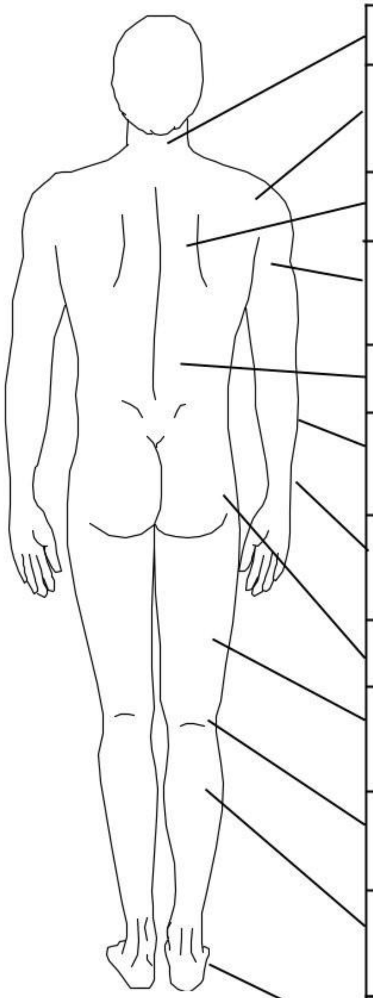

© Cornell University, 2003

| Body part | | During the last work <u>week</u> how often did you experience ache, pain, discomfort in: | | | | | If you experienced ache, pain, discomfort, how uncomfortable was this? | | | If you experienced ache, pain, discomfort, did this interfere with your ability to work? | | |
|---|---|---|---|---|---|---|---|---|---|---|---|---|
| | | Never | 1-2 times last week | 3-4 times last week | Once every day | Several times every day | Slightly uncomfortable | Moderately uncomfortable | Very uncomfortable | Not at all | Slightly interfered | Substantially interfered |
| Neck | | ☐ | ☐ | ☐ | ☐ | ☐ | ☐ | ☐ | ☐ | ☐ | ☐ | ☐ |
| Shoulder | (Right) | ☐ | ☐ | ☐ | ☐ | ☐ | ☐ | ☐ | ☐ | ☐ | ☐ | ☐ |
| | (Left) | ☐ | ☐ | ☐ | ☐ | ☐ | ☐ | ☐ | ☐ | ☐ | ☐ | ☐ |
| Upper Back | | ☐ | ☐ | ☐ | ☐ | ☐ | ☐ | ☐ | ☐ | ☐ | ☐ | ☐ |
| Upper Arm | (Right) | ☐ | ☐ | ☐ | ☐ | ☐ | ☐ | ☐ | ☐ | ☐ | ☐ | ☐ |
| | (Left) | ☐ | ☐ | ☐ | ☐ | ☐ | ☐ | ☐ | ☐ | ☐ | ☐ | ☐ |
| Lower Back | | ☐ | ☐ | ☐ | ☐ | ☐ | ☐ | ☐ | ☐ | ☐ | ☐ | ☐ |
| Forearm | (Right) | ☐ | ☐ | ☐ | ☐ | ☐ | ☐ | ☐ | ☐ | ☐ | ☐ | ☐ |
| | (Left) | ☐ | ☐ | ☐ | ☐ | ☐ | ☐ | ☐ | ☐ | ☐ | ☐ | ☐ |
| Wrist | (Right) | ☐ | ☐ | ☐ | ☐ | ☐ | ☐ | ☐ | ☐ | ☐ | ☐ | ☐ |
| | (Left) | ☐ | ☐ | ☐ | ☐ | ☐ | ☐ | ☐ | ☐ | ☐ | ☐ | ☐ |
| Hip/Buttocks | | ☐ | ☐ | ☐ | ☐ | ☐ | ☐ | ☐ | ☐ | ☐ | ☐ | ☐ |
| Thigh | (Right) | ☐ | ☐ | ☐ | ☐ | ☐ | ☐ | ☐ | ☐ | ☐ | ☐ | ☐ |
| | (Left) | ☐ | ☐ | ☐ | ☐ | ☐ | ☐ | ☐ | ☐ | ☐ | ☐ | ☐ |
| Knee | (Right) | ☐ | ☐ | ☐ | ☐ | ☐ | ☐ | ☐ | ☐ | ☐ | ☐ | ☐ |
| | (Left) | ☐ | ☐ | ☐ | ☐ | ☐ | ☐ | ☐ | ☐ | ☐ | ☐ | ☐ |
| Lower Leg | (Right) | ☐ | ☐ | ☐ | ☐ | ☐ | ☐ | ☐ | ☐ | ☐ | ☐ | ☐ |
| | (Left) | ☐ | ☐ | ☐ | ☐ | ☐ | ☐ | ☐ | ☐ | ☐ | ☐ | ☐ |
| Foot | (Right) | ☐ | ☐ | ☐ | ☐ | ☐ | ☐ | ☐ | ☐ | ☐ | ☐ | ☐ |
| | (Left) | ☐ | ☐ | ☐ | ☐ | ☐ | ☐ | ☐ | ☐ | ☐ | ☐ | ☐ |
